# Quality Improvement in Clinical Research: ESA Utilization to improve Patient Safety Monitoring, time management, and Research Compliance

**DOI:** 10.1101/2021.11.05.21265341

**Authors:** Maria Del Pino, Ana Bonmati, Patricia Mendoza

## Abstract

We analyzed the capabilities of Electronic Signature Administrators (ESA) in reporting adverse and serious adverse events (AE/SAE) in clinical trials to improve Principal Investigator (PI) oversight. The current and most conventional way to sing on AE/SAE reports, by wet ink paper signatures, faces long report completion times, miscommunications, and a high rate of error that can put the study and the patient at risk. We demonstrate here that ESAs can easily take over this task, speeding up the process and enhancing patient’s safety and research compliance. Our results show that the average of 31 days to obtain wet-signatures on AE/SAE reports is reduced to merely 3 days after the implementation of ESA. We also found that research professionals spend more than half an hour in commuting each time they need to get PI signatures, while this time is zero if ESAs are used. Finally, an anonymous survey distributed to coordinators shows that ESAs can be well received when implemented since 86% have used an ESA and are satisfied with it or are interested in using it, while only 14% of the participants are either not interested or not satisfied.

## Introduction

Clinical trials are essential for the improvement and development of current and new treatments as well as new ways to detect, diagnose, and assess diseases with the purpose of prolonging the patients’ lifetime and their quality of life [1]. Although conducting a clinical trial may appear straightforward, the management is complex, lengthy and expensive [2,3]. In some cases, up to 40% of the cost to bring a new treatment to the market is related to paper-based processes [4]. In fact, in clinical research there are many non-essential activities that increase the challenge in conducting a trial and delays its initiation, and therefore, delays the access to new treatments for patients. Some of these activities, which are constantly increasing, include different policies, tests, regulations, protocols, etc. While most of these tasks are indeed needed for patient safety and control, they can produce an overwhelming situation to clinical researchers, especially due to the lack of innovation to help them handle all the paperwork.

Despite all these documentation and regulatory burden to conduct clinical trials [5], research professionals are committed to apply high quality standards for the development of new treatments. Their work is especially important in the early stages of the clinical research studies, when new medical products could lead to undesired health conditions in patients. For the purpose of this work, we distinguish two undesired conditions: Adverse Events and Serious Adverse Events. An Adverse Event (AE) is defined as any unfavorable and unintended, symptom, or disease temporally associated with the use of an investigational medical product, whether considered related to the product or not. If the adverse event is important enough to lead to severe complications, threatening the life or function of the person then, it is considered a Serious Adverse Event (SAE) [6].

Some of the AEs and SAEs can be critical for patients and require immediate reporting to regulatory entities [7]. Therefore, these events must be closely tracked by the Principal Investigator (PI) in a timely manner. The PI is the research professional legally responsible for conducting the study and accountable for the supervision of the tasks performed by other research and professionals, regardless of the qualifications and expertise they may have [8]. To prove PI active participation in the study, he/she has to sign and date research related documents. An effective and accurate PI oversight and adverse event adjudication is key to guaranteeing patient safety, while being compliant with the current regulation and institutional policies, especially when a potential AE/SAE arises. In this case, the PI conducts a timely assessment of the event, stablishing the relationship/causality, severity, seriousness and expectedness of the AE/SAE. In addition, when this happens, the PI is responsible for reporting the pertinent information and documentation to both, the sponsor and the IRB (Institutional Review Board) within the period of time specified in the ICH GCP E2A guideline [4]. Therefore, the assessment and reporting of adverse events are time sensitive matters. To accomplish this task, the clinical trial coordinators are essential members of the research team. However, due to the multiple demands that these research professionals are exposed every day [7,8] can be very difficult to obtain signatures on the participant’s medical report and/or other research documents in a timely manner. Very often, it takes long time for the investigator to review the documents, sign, and date them as a proof of their timely and active participation in the research process. In fact, we don’t consider that the PI is fully aware of the AE/SAE until he/she reads, sign and date the associated medical report, which includes all the details of the event.

To facilitate PI oversight, while being compliant with regulations [4], research professionals usually send an email to the investigator to alert him/her about the potential AE/SAE, in a timely manner and ask to adjudicate the event based on the description provided in the email. However, this way of communication is not always complete and fails to provide essential information that the investigator needs to properly adjudicate the event, such as medical history of the patient, current medication, symptoms associated with AE/SAE, etc., which can lead to misunderstandings and may interfere with the proper assessment and adjudication of the adverse event. Another way of communication used is by leaving the research documents on the desk of the PI’s office, expecting that he or she will review and wet-sign the documents associated with a potential AE/SAE in a timely manner. Nevertheless, this way of obtaining wet-signatures is of a serious concern from a regulatory standpoint, because documents can sit on desks for several days or even get lost, putting the study and the participant at risk.

The ideal scenario is when the investigators can adjudicate potential AE/SAE directly on patient’s records, where all the information needed to assess the new event is documented. This method would prevent miscommunications, errors and the incorrect adjudications of potential adverse events. A solution to approach to the ideal situation is by using an Electronic Signature Administrators (ESA). An ESA is an electronic application that enables people to send, sign, and approve documents anytime from anywhere, by using only an electronic device and a net connection. However, the use of this technology its being adopted very slowly due to some concerns related to cost, regulatory compliance, and its implementation. This tool has already demonstrated the expedition of procedures, saving staff time, and a highly reduction of errors on diverse research documents [10].

## Methods

This is a single-institution, nonrandomized prospective and comparative process improvement project that analyses the time to obtain PI signatures on medical records associated with potential AE/SAE before and after implementing an ESA. For this project, ESAs have only been used after being approved by the sponsor, institution, the IRB, the clinical trial manager, and the director of the department where the tool was implemented.

To conduct this study, we used DocuSign®, [11] and EPIC, our electronic medical record system [12] as ESAs. We chose these systems for their capabilities regarding security, accessibility, data integrity, and their adherence to institutional policies and certifications.

We calculated the average time collecting PI wet signatures in participant’s medical reports related to potential AE/SAEs in a Research Institute for 1 year (May 2019 to May 2020). During this period 28 different studies were carried out and a total of 694 AE/SAE were reported. We applied the statistical sample size formula (Equation 1) to choose a more manageable size of the population while keeping high accuracy. The formula allows us to determine how many reports we need to consider to reflect the target population as precisely as needed.

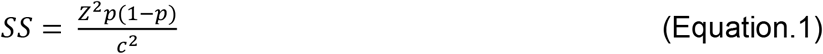

In Eq. 1, we used *Z* = 1.96, which corresponds to the tabulated value of considering 95% of confidence level, and *c* = 0.07, which represents the confidence interval or margin error of 7%. *p* is the estimator of the proportion, which was set to 0.5, corresponding the maximum variance of a binomial distribution. Then we applied the correction for finite population (Equation 2)

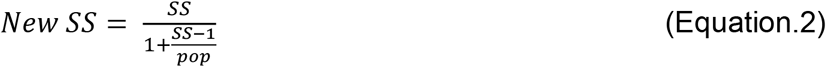

where *pop* is the 694 adverse events reported in a year. By using both equations we reduced our population to a sample size of 153 adverse events, which were chosen randomly. On the other hand, we implemented the ESA in 9 different studies, where 76 potential AE/SAE were notified for 1 year (from Aug 2020 to Aug 2021). For each one of these we obtained the date when the research coordinator was notified or found the AE/SAE and the date when PI signed it on the patient’s medical report.

We also assessed the time that a research professional spends in collecting PI wet signatures on documents related to clinical trials, which includes commuting to different areas of the hospital, the waiting time for the PI to be available to wet-sign the documents, and the time clarifying the reviewed entities and signing. For this purpose, we distributed questionnaires to the research professionals within 2 days of collecting their signatures. We collected 23 anonymous questionnaires (from March 2021 to May 2021). We want to highlight here that the data was treated to eliminate outliers that considered an excessive spending time. For instance, we found questionnaires were the commuting time were over 2h. We considered this an excessive amount of time to commute from one area of the hospital to another. As a result, we reduced these commuting times to a maximum of 30 minutes. Nevertheless, we have cases where some research professionals, that work in external branches of the Research Institute, may need to commute to the main facility to collect PI signatures. In these situations, professionals spend a large amount of time to collect wet-signatures on research documents.

Finally, we also considered important to capture the opinion of research professionals about the use or interest of using ESAs to report AE/SAEs. To obtain such information, we distributed questionnaires to the research coordinators. A total of 44 anonymous surveys were collected in several research departments across the institution. On this questionnaire, we asked to the professionals whether they had previous experience using an ESA. If the answer was “yes”, they had to rate this tool between 1 and 10 on several features: convenience, easiness, speed and, if based on their opinion, it could help them to provide a better service to sponsor and patients. However, if the research professional did not have experience with ESA, they had to explain the reasons and if would like to try it or not. The results obtained provided an interesting insight into the clinical research community to decide about an ESA implementation in the institution.

## Results

We found that it takes an average of 32 days to obtain evidence of PI oversight wet-signatures in the patient’s medical records when ESA is not implemented. The range goes from 0 to more than 300 days in many occasions. This amount of time is surprisingly high for a patient’s safety related matter. As it can be seen in Figure.1, in 37% of the cases it took 5 or less days to obtain PI signatures. We consider this window of time the maximum admissible to complete this task. However, most of the wet-signatures were obtained outside of this window. Specifically, 63% of the AE/SAE reported events obtained PI signatures within more than 5 days. In 37 out of the 153 studied cases, the clinical coordinator was able to obtain the signatures 3 weeks after the day the AE/SAE was reported. These results, clearly contrast with the ones obtained after implementing ESA (see figure 2). We found that the 32 days on average to obtain signatures and PI adjudication were highly reduced to 3 days on average when utilizing an ESA. The range of days spent for signing using ESA was also reduced and goes from 0 to 34 (where 34 was an outlier that was included in the analysis).

**Figure.1.**
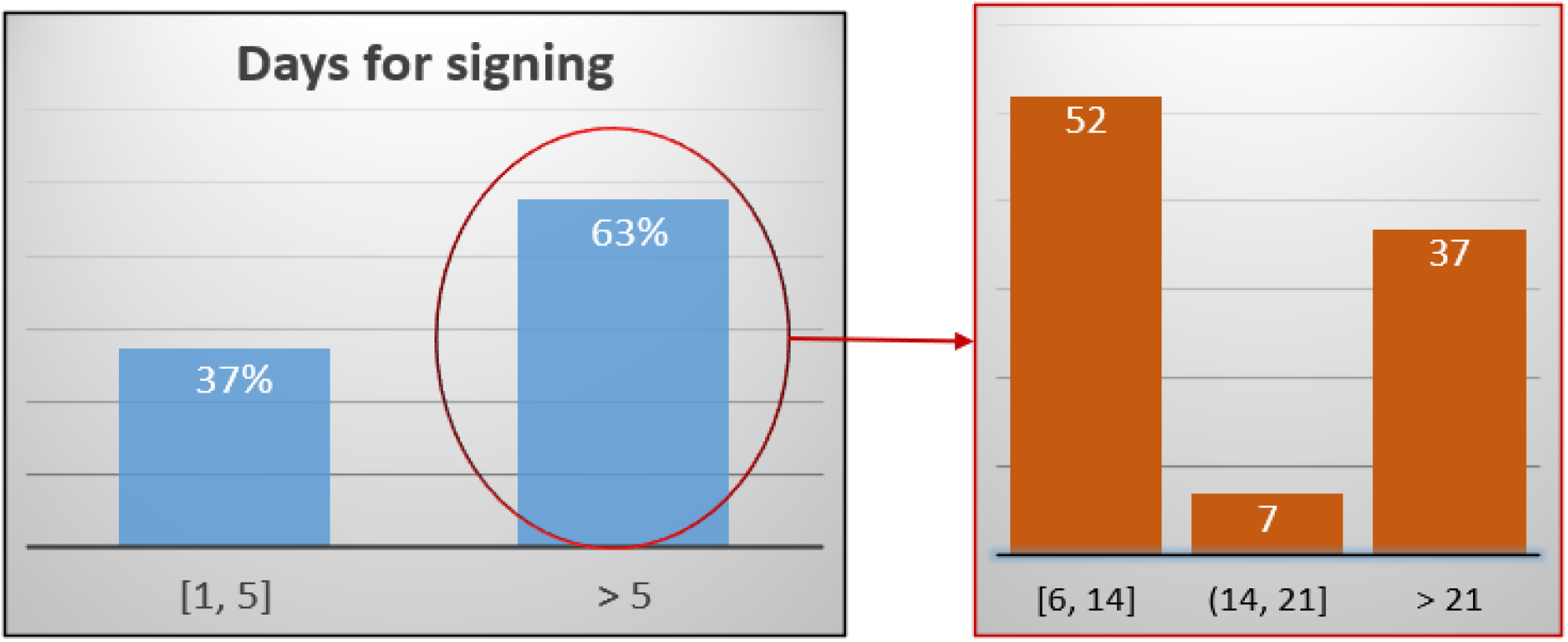
Total waiting time (in days) to collect PI signatures. Left – Percentage of events signed within 5 days or more from the day that AE/SAE was reported. Right – Amount of events reported in more than 5 days broken down per weeks

**Figure 2.**
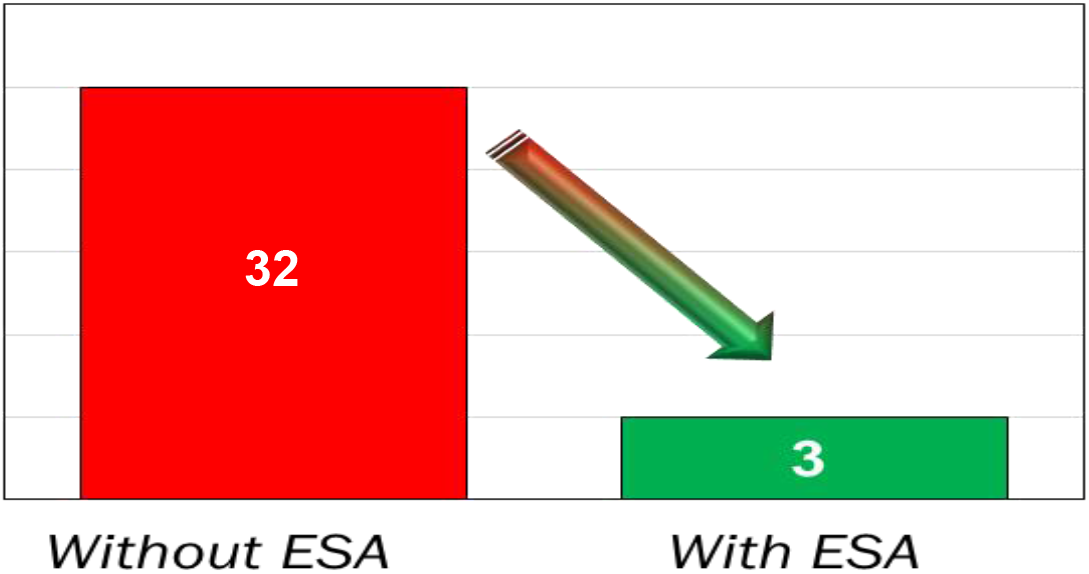
Average of days to report AE/SAE without and with using an ESA

Keep in mind that most sponsors want AE and SAEs reported within 24 hours, the FDA asks for critical safety information as soon as possible or at least within 15 calendar days the sponsor received the report and IRBs have their due dates [6].

A solution to reduce time collecting signatures without ESA implementation, would be that research coordinators going to the PI’s office more often. However, this is a continuous hassle and can be a waste of time for the research professionals, decreasing employee satisfaction. We found that research professionals spend 37 minutes on average each time they go to the PI’s office to obtain wet-signatures. In some cases, this time goes to over an hour. The average time for commuting to other areas to the hospital is 16 minutes. The waiting time for the PI to be available to review and wet-sign the documents takes an average of 9 minutes and there is an average of 12 minutes to actually obtain the signatures. The median obtained for the entire process of reaching the PI and getting the signatures is 31 minutes (see Figure 3 for more information). Interestingly, in almost 20% of the cases the research coordinator had to come back to the PI’s office because not all of the signatures were collected. In contrast, the time spent in commuting to collect signatures using an ESA is obviously zero. Less time spent collecting wet-signatures implies more times focusing on patient’s care and study goals.

**Figure. 3.**
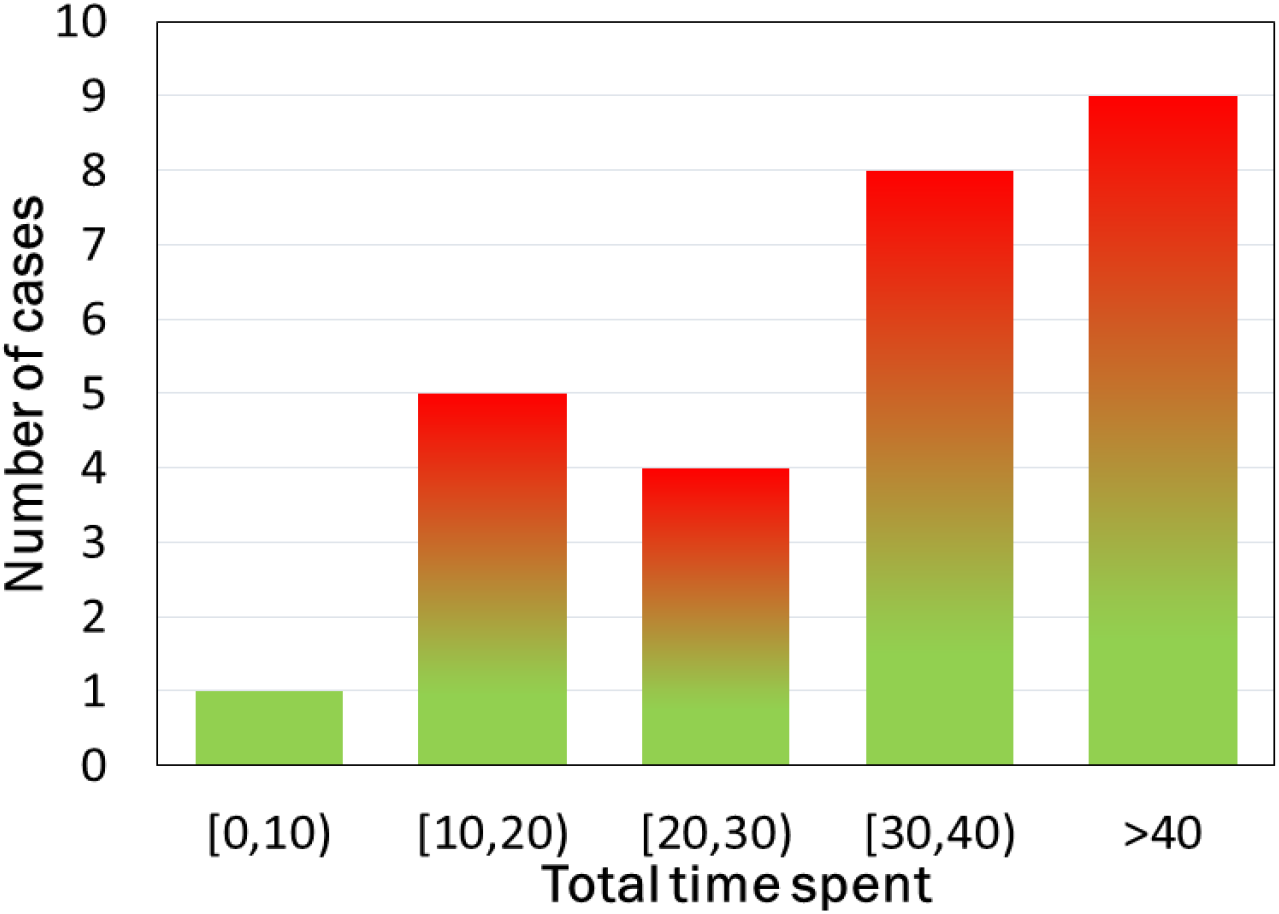
Commuting time (in minutes) to collect PI signatures. Average is 37 minutes

Finally, we found that 59% of the research professionals who have used ESA have a positive feedback. They scored with more than 8 in all the questions. We also observed a 7% negative feedback from research professionals that did not like the ESA; they scored less than 6 in all the questions. In addition, we learned that 27% of the research professionals have never used this tool but are interested in trying it. However, 7% of the research professionals have neither tried this tool and not interested in use it. The interesting takeaway of this study is that 86% of professionals have used it and like it, or are interested in using it (Table 1)

**Table 1.**
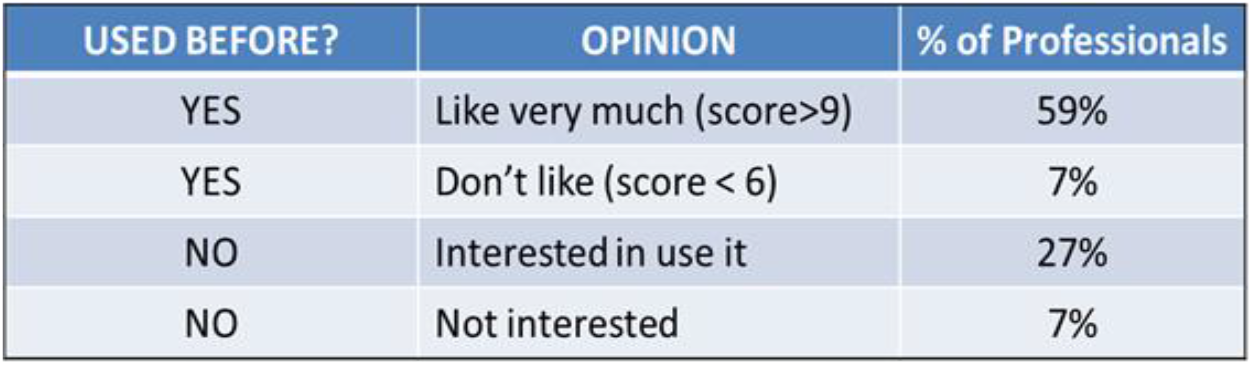
Results of the opinions obtained from research coordinators regarding the use of an ESA.

| USED BEFORE? | OPINION | % of Professionals |
| --- | --- | --- |
| YES | Like very much (score>9) | 59% |
| YES | Don't like (score < 6) | 7% |
| NO | Interested in use it | 27% |
| NO | Not interested | 7% |

## Discussion

The implementation of ESA in clinical research has never been more important than nowadays, facing the COVID 19 pandemic. The coronavirus extended across the globe affecting our hospitals and health systems, which forced many professionals to work remotely and conduct many research related activities from distance [10]. The pandemic has made urgent for clinical research professionals to innovate and to shift from paper processes to electronic formats [13]. However, its implementation remains slow [14].

The results obtained from this process improvement study demonstrates that by implementing an ESA we drastically decrease the time of collecting PI signatures versus the traditional way. In addition, an ESA greatly improves communication among research professionals since the PI signs and dates directly on the patient medical records, avoiding miscommunication and inaccurate information, thus facilitating proper adjudications of AE/SAE. In addition, ESA also prevent common errors related to missed or wrong dates, since they require the signer to complete all the fields to proceed.

ESA can also enhance the productivity of research professionals and accelerate processes because the documents can be prepared and signed remotely, thanks to the institutional security in place and encrypted password. Therefore, no more commuting times between offices or departments for coordinators, no more waste of time waiting for PI’s availability or reviewing documentation. This automates workflows and facilitate investigator oversight from any location, which leads to saving time and money while enhancing the experience of our customers and patients, who will be pleased to see all the research documentation reviewed and signed in a timely manner. Furthermore, while traditionally research documents are difficult to control, the use of ESA allows to easily see which documents have been signed and which are pending. Then, it sends reminders to prevent missed documents due to a human error.

This software provides high availability, fault tolerance and treat isolation due to the safety security and standards. It is very safe because of the documentation encryption, as well as its adherence to policies and certifications. ESA saves copies of the documents in internal repositories so, they are ready for review and inspect at any time. Despite all the ESA advantages, its implementation requires a significant initial investment, which usually includes software fees, development of SOPs and workflows. Other challenges can also arise, such as electronic security, unfamiliarity with electronic devices and organizational inertia. Migration from wet-signature to electronic signature may be confusing at the beginning and HIPAA concerns with sending data. However, the results obtained in the survey distributed permit allow us to assume that the ESA familiarization can be smoother than anticipated and very beneficial for the coordinators. In the future, a multisite study could shed more light in terms of cost efficiency, profitability and the possible challenges that can arise when implementing an ESA. Finally, ESA can also be useful in taking over other tasks in clinical research such as patients consenting, regulatory documents, etc.

## Conclusion

The results of this single-site study demonstrate that the use of ESA can ease and accelerate the reporting and adjudication of adverse events in clinical trials by speeding up PI oversight while improving participant’s safety. The results obtained in this evidence-based process improvement project show how the average time to obtain PI oversight wet-signatures is highly reduced from 32 to 3 days. In addition, ESA enables more efficient AE/SEA adjudication, preventing possible miscommunications and inaccurate data, and contributes with stronger research compliance, saving time to the professionals and therefore, saving money to the institution. Furthermore, the tool is secure, efficient and accessible, which will contribute to enhance the satisfactions of sponsors, patients and professionals. These great advantages contribute with innovation for the development of new treatments and, therefore, support the wellbeing of our society.

## Data Availability

Questionnaires
Information about Principal Investigator oversight on Adverse events before and after the implementation of an Electronic Signature system

